# *Ex Vivo* Phosphorus-31 Solid State Magnetic Resonance Spectroscopy Identifies Compositional Differences Between Bone Mineral and Calcified Vascular Tissues

**DOI:** 10.1101/2025.09.27.25336805

**Authors:** Jingting Yao, Christian T. Farrar, Elena Aikawa, David E. Sosnovik, Brianna F. Moon, Aditi Kulkarni, Jerome L. Ackerman

## Abstract

**Aim:** Physiological bone mineralization and ectopic vascular calcification share similarities in the composition of calcium phosphate minerals. Evidence suggests a connection between the underlying biological mechanisms driving the deposition of bone mineral and cardiovascular calcification. Therefore, understanding the chemistry and composition of bone mineral and vascular calcification may be important for the development of effective treatments and diagnostic tools for cardiovascular diseases, as pharmacological interventions for the treatment of one process might affect the other. The goal of this study was to identify and compare compositional features of calcium phosphates in bone and calcified vascular tissues using phosphorus-31 (^31^P) solid state cross-polarization (CP) magic angle spinning magnetic resonance (MR) spectroscopy, a specialized technique that provides compositional information unattainable through conventional chemical analysis.

**Methods:** Solid state MR spectra were acquired from biological specimens of human trabecular bone (n = 1), human vascular plaque (n = 1), human calcified aortic valves (n = 5), as well as calcified aortic tissues of apolipoprotein E-deficient mice (n = 1) fed a high cholesterol diet. Synthetic hydroxyapatite (Ca_10_ (OH_2_) (PO_4_)_6_) and synthetic brushite (CaHPO_4_ · 2H_2_O) were used to model the solid state ^31^P MR spectra of the phosphate ion PO_4_^−3^ and hydrogen phosphate ion HPO _4_^−2^, respectively. Qualitative spectral features and quantitative metrics derived using Herzfeld-Berger analysis were assessed to characterize mineral composition and maturity.

**Results:** Solid state ^31^P MR spectra of all human specimens studied suggested a well-ordered crystal structure dominated by unprotonated phosphate (PO_4_^−3^), consistent with mature bone-like mineral. These specimens exhibited long CP time constants (700–900□µs) and modest chemical shift anisotropy. In contrast, the calcified mouse aorta spectrum showed pronounced sidebands, a short CP time constant (~270□µs), and a more prominent HPO_4_^−2^ component—features indicative of immature, newly deposited mineral.

**Conclusion:** ^31^P solid state MR spectroscopy reveals differences in the phosphate and hydrogen phosphate ion content among the calcified tissues studied. This technique could potentially be an important complement to basic studies of pathological calcification in atherosclerosis and related calcific disorders.

## 1 Introduction

Physiological bone mineralization and pathological vascular calcification exhibit some similarities in their underlying mechanisms and calcium phosphate mineral composition (1, 2). Although the two processes are physiologically distinct, they share some cellular pathways and molecular mediators, including common proteins and regulatory factors (3–9). Inflammation and macrophage activities are key drivers of vascular calcification, and osteoclasts—the bone-resorbing cells—originate from the same hematopoietic lineage as macrophages (10, 11). Epidemiological studies have reported associations between osteoporosis and increased cardiovascular risk (12). Clinical observations further support this biological linkage: pharmacological agents used to treat cardiovascular diseases (CVDs) can influence bone metabolism, and conversely, treatments for osteoporosis may affect vascular calcification (13–15). Specifically, bisphosphonates (BPs), the most common anti-osteoporotic agents, inhibit bone resorption by binding to hydroxyapatite and suppressing osteoclast activity (16). BPs are non-hydrolyzable analogs of inorganic pyrophosphate, an endogenous inhibitor of vascular calcification (17, 18). BPs have been linked to altered lipid profiles and reduced aortic calcification (19). Moreover, statins have shown associations with increased bone mineral density [15]. Estrogen deficiency, which accelerates bone loss, is also a known risk factor for atherosclerosis and vascular calcification (20, 21).

Vascular calcification is a key feature of atherosclerosis and a well-established predictor of cardiovascular events. While coronary artery calcium scores reflect total calcium burden, studies suggest that the chemical composition and structural organization of calcium deposits may more directly influence plaque stability and clinical outcomes (22–28). Beyond serving as a risk marker, mineral composition may reflect distinct stages of plaque development with differing biological implications, potentially clarifying mechanisms of disease and inform the design of targeted therapies aimed at specific phases of calcification.

In vertebrates, the final mineral product of biomineralization is a poorly crystalline, nonstoichiometric form of hydroxyapatite (Ca_10_(OH)_2_(PO_4_)_6_), often referred to as biological apatite. Solid state magnetic resonance (MR) spectroscopy evidence suggests that mature biological apatite may evolve from an initial precursor resembling dicalcium phosphate dihydrate (CaHPO_4_·2H_2_O, brushite), which is deposited in soft tissue during the earliest stages of mineralization (29). Alternative models of vertebrate mineralization propose other precursors—such as amorphous calcium phosphate (Ca_3_(PO4)_2_) or octacalcium phosphate (Ca_8_(HPO_4_)_2_(PO_4_)_4_·5H_2_O)—that may also transform into mature apatite over time. Hydroxyapatite, containing unprotonated phosphate (PO_4_^−3^), and brushite, containing protonated phosphate (HPO_4_ ^−2^), have been used as chemical models for the respective unprotonated and protonated phosphate ions in biological calcification. Therefore, one measure of the maturity of the biological mineral may be obtained by assessing the relative concentrations of HPO_4_^−2^ and PO_4_^−2^, given that mature bone mineral is dominated by PO_4_^−3^ and has a lower HPO_4_ ^−2^ concentration (29, 30).

No known study has yet differentiated and quantified phosphate ion species in calcified vascular tissues, and such compositional information is unattainable using conventional chemical analysis. Clinical evaluations using cardiac computed tomography (CT), intravascular ultrasound and optical coherence tomography present structural details with qualitative compositional information (31–37). *Ex vivo* evaluations of plaque and calcification include electron microscopy and micro-CT (µCT) for morphological measurements, and histology and immunohistochemistry for cellular examination. All these methods are unable to distinguish specific phosphate ion types or provide quantitative compositional information. Direct chemical analysis, as well as techniques like Fourier transform infrared spectroscopy and X-ray diffraction encounter significant challenges due to interference from residual water and soft-tissue constituents. Solid state MR spectroscopy thus offers a valuable alternative capable of identifying and quantifying phosphate ion species in mineralized tissues.

In this study, we employed two fundamental solid state MR spectroscopy techniques to analyze calcified tissue specimens: magic angle spinning (MAS) and cross-polarization (CP). The more common type of MR spectroscopy is performed on fluids, such as soft tissues *in vivo* or chemical solutions, where the signals from individual chemical groups (e.g., methyl groups, sodium ions, or water molecules) appear as resonance lines characterized primarily by their positions along the horizontal axis, known as the chemical shift, and measured in parts per million (ppm). Each chemical shift value uniquely corresponds to the identity of the respective chemical group. In contrast to fluids, in which rapid molecular tumbling and diffusion occur, molecules within solids are fixed in positions and orientations, making their chemical shifts orientation dependent. Consequently, MR spectra of solid substances, such as calcium phosphate mineral crystals present in calcified tissues, exhibit broad resonance patterns spanning a wide frequency range, complicating direct identification of chemical groups from single-valued chemical shifts. Such broad resonance profiles, known as powder patterns, reflect the geometric symmetry of the chemical group or ion. The central position of the powder pattern corresponds to the isotropic chemical shift that would be observed if the molecules were freely mobile, as in a fluid state.

MAS facilitates MR spectroscopy of solids by rapidly rotating the specimen around an axis inclined at approximately 54.7 degrees relative to the external magnetic field, the so-called “magic angle” (38). Rapid rotation partially averages orientation-dependent chemical shifts, analogous to random molecular tumbling in liquids, significantly reducing the breadth of powder patterns toward narrower resonance lines. To achieve a single-valued chemical shift, rotation rates must surpass the intrinsic width of the powder pattern, a condition that is technically challenging. Typically, lower rotation rates result in partial narrowing, producing a pattern characterized by sidebands spaced at intervals equal to the rotation frequency (e.g., Fig. 1).

**Figure 1.**
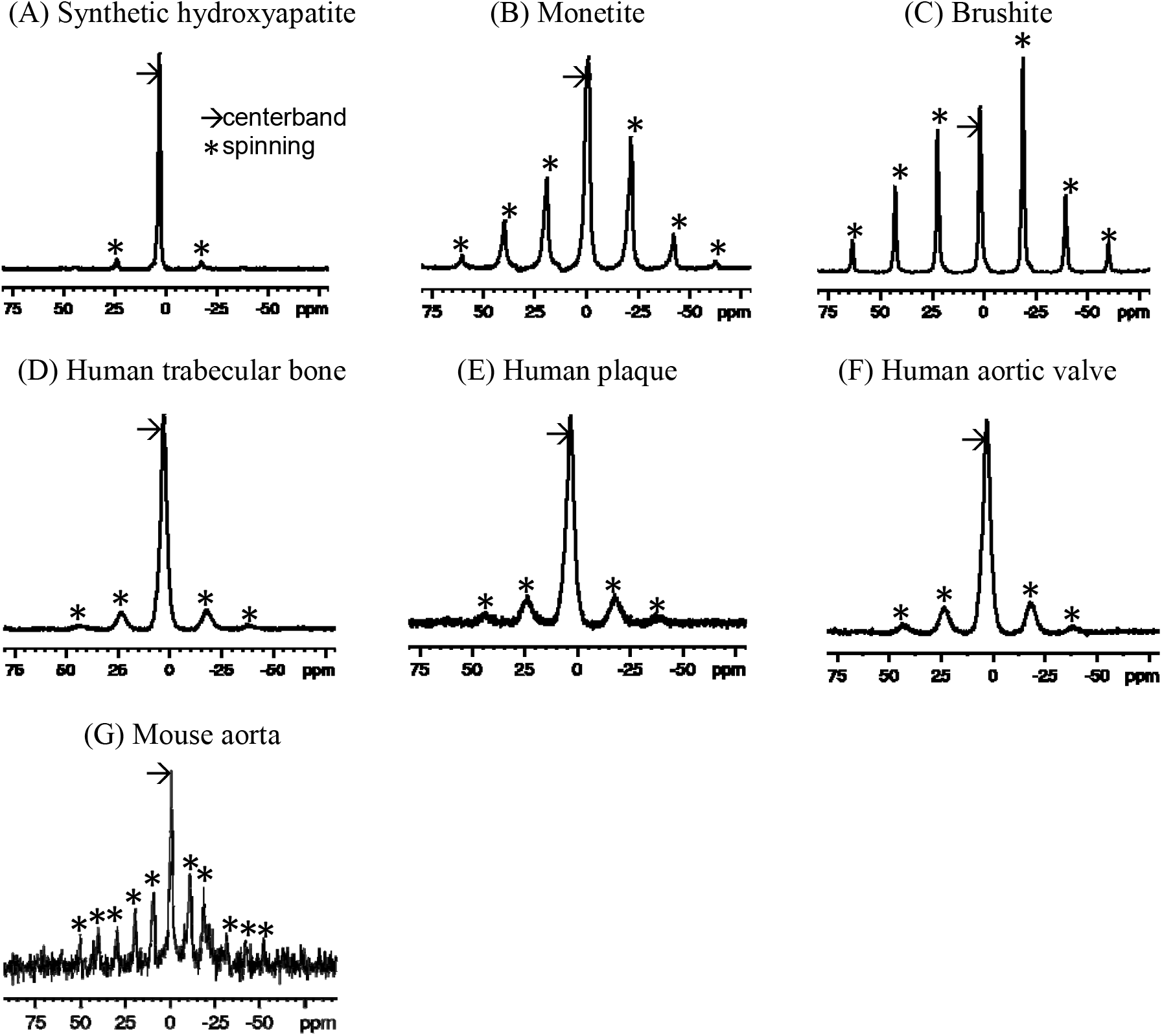
^31^P single-pulse MAS spectra of synthetic calcium phosphate compounds (A-C) and ^1^H-^31^P CPMAS spectra of mineralized tissues (D-G). The spin rate for (A-F) was 5 kHz (~21 ppm), while that for (G) was 2.5 kHz (~10.5 ppm). Note that sidebands (*) are spaced at the spin rate. Chemical shifts are referenced to 85% H_3_PO_4_.

CP serves two main functions in solid state MR spectroscopy. First, CP can enhance signals from nuclear isotopes with inherently low signal-to-noise ratio (SNR), such as those with low MR frequencies, low isotopic or chemical abundances, or long spin-lattice relaxation times T_1_. This enhancement is achieved by transferring magnetization from isotopes with higher SNR (commonly protons, ^1^H) to the isotope of interest through simultaneous radiofrequency (RF) excitation. Second, CP is useful for spectral editing, differentiating chemical groups based on their proximity to the isotope source of magnetization. CP and MAS are frequently combined as cross-polarization magic angle spinning (CPMAS). It is important to note that CP magnetization transfer and the magnetization transfer effect in clinical MRI are fundamentally different. The former occurs via transverse magnetization exchange on microsecond-to-millisecond time scales, whereas the latter occurs on significantly longer (millisecond-to-second) time scales as a result of the diffusion of magnetization transported either by chemical diffusion of nuclear spins or by spin-spin coupling.

To date, aside from straightforward non-CP phosphorus-31 (^31^P) MAS measurements (39, 40), solid state MR techniques have not been utilized for examining vascular calcification. ^1^H-^31^P solid state CPMAS MR spectroscopy has proven effective in quantifying bone mineral and solid organic matrix *ex vivo* (41–47). In biological specimens, distinguishing phosphate ions such as PO_4_^−3^ and HPO_4_^−2^ based on chemical shifts (3.1 ppm and 1.4 ppm, respectively) may be challenging, as inherent crystallographic disorder leads to broad resonance linewidths even with MAS. Thus, measurement of CP transfer rates, depend strongly on the proton-phosphorus distances, may help discriminate between these ions. For instance, rapid CP occurs in HPO_4_^−2^ ions in which ^1^H and ^31^P nuclei are spaced by only a few tenths of a nanometer. Conversely, hydroxyapatite consists of spatially separated OH^−^ ions (protons) and PO_4_^−3^ ions (phosphorus), leading to slower CP. Thus, the CP rate is reflective of the local crystal structure and can inform about the types of phosphate ions present, as established by prior studies in bone (43, 48). Indeed, ^1^H-^31^P CPMAS investigations of progressively mineralizing embryonic to mature chicken bone have demonstrated a clear reduction in HPO_4_^−2^ content from initial embryonic mineralization (approximately 8 days post-egg laying) to fully mature bone in 75-week-old chickens (30, 43). Given the biological parallels between bone mineralization and vascular calcification, we hypothesize that CPMAS could similarly reveal compositional variations within calcified vascular tissues, potentially providing insights into the mineral maturity of these tissues.

In this study, we applied CPMAS MR spectroscopy to investigate pathological calcifications from human plaque, human aortic valve, and the aorta of apolipoprotein E (ApoE)-deficient mice, along with human trabecular bone specimens. To characterize mineral maturity, we compared these spectra to reference spectra of calcium phosphates commonly used as mineral models of biological calcification. CP transfer rates were derived to assess differences in crystal structure and composition. Additionally, we applied Herzfeld-Berger (HB) analysis (49) to extract the principal values of the chemical shift tensor (CST), parameters mathematically modelling the orientation-dependent variation in chemical shifts, from the spectral data. Given adequate spectral quality, indicated by clear identification and assignment of spinning sidebands to phosphate ions (PO_4_^−3^ and HPO_4_^−2^), HB analysis can reliably determine both the relative abundance of these phosphate species and their respective chemical shift tensors, despite significant spectral overlap.

## 2 Materials and methods

### 2.1 Specimens

Commercial synthetic compounds included hydroxyapatite Ca_10_(OH)_2_(PO_4_)_6_, brushite (CaHPO_4_·2H_2_O), and monetite (CaHPO_4_) (Fisher Scientific, Hampton, NH, USA) were used without further purification or analysis. Human specimens included trabecular bone from autopsy (n = 1), calcified aortic plaque (n = 1) from autopsy, and calcified human aortic valves (n = 5) acquired at the time of surgical excision and placed in neutral buffered formalin. Calcified aortic tissue (aortic root and arch vessels) was harvested en bloc from 60-week-old ApoE-deficient mice (n = 1), that had been fed an atherogenic diet from 10 weeks of age; mouse specimens were preserved in formaldehyde buffer. All specimens were stored at –80 °C until MR scanning. Although specimens from two ApoE-deficient mice were obtained, only one yielded a spectrum with sufficient signal-to-noise ratio (SNR) for analysis. The second mouse specimen was excluded due to unstable spinning and low SNR, which precluded reliable spectral fitting.

Prior to MR spectroscopy, intravital fluorescence microscopy of the carotid artery in these mice was performed using a bisphosphonate-derived imaging agent (OsteoSense 750; dose 2 nmol/150 μL). µCT of the mice was also performed to detect calcification.

Animal studies were approved by the respective Institutional Animal Care and Use Committees. Use of discarded human specimens was approved by the Massachusetts General Hospital Institutional Review Board.

### 2.2 MR spectroscopy

Spectra were acquired using a Bruker Biospin (Billerica, MA, USA) Avance III multinuclear MR spectrometer equipped with a 14 Tesla Magnex Scientific (Oxford, UK) wide bore magnet (f_1H_ = 600 MHz and f_31P_ = 243 MHz). Samples, either compounds or calcified portions of the biological specimens, were tightly packed into 4 mm zirconia MAS rotors with a volume of about 78 μL. Both single-pulse ^31^P MAS and ^1^H-^31^P CPMAS spectra were acquired. For CP acquisitions, the 90° RF pulse length was 4.5 µs. (B_1_ field 55 kHz) on both ^1^H and ^31^P channels. Single-pulse ^31^P spectra used a 90° RF pulse of 4.1 µs. The MAS spin rate was 5 kHz for most specimens, with 32 averages per spectrum and 2 s recycle delay (TR), resulting in 64 s acquisition duration per spectrum. For mouse aortic samples, which had lower mineral content and showed spinning instability due to tissue heterogeneity, the MAS rate was reduced to 2.5□kHz. To compensate, 7,400 scans were acquired per spectrum, giving a total acquisition time of approximately 4.1□hours to ensure adequate signal-to-noise ratio (SNR). CP spectra were also acquired with varying CP times to measure ^1^H-^31^P CP time constants (*τ*_CP_) and ^1^H rotating frame spin-lattice relaxation time constants (*T*_1 *ρ*_) for the buildup and loss of magnetization, respectively.

### 2.3 MR data analysis

Spectral data were processed offline using Bruker TopSpin 3.6.2 (Bruker, Billerica, MA, USA). Up to 2 ppm line broadening was applied to the mouse aorta spectra for reasonable SNR. For each series of CP spectra with varying CP times, the spectrum with the most intense centerband exhibited the highest SNR, and its phase correction was applied to all spectra in the series. Because it is typically not possible to use an internal chemical shift reference in solid state MR spectroscopy, a hydroxyapatite external reference set to 3.1 ppm (corresponding to 85% phosphoric acid defined as 0 ppm) was applied.

Herzfeld-Berger analysis was performed on the spectra used for phasing, employing the software tool ssNake (50) to extract the CST. A CST is characterized by three numbers which can be mathematically expressed in multiple ways: as three principal values *δ*_11_, *δ*_22_, and *δ*_33_ where *δ*_11_ ≥ *δ*_22_ ≥ *δ*_33_, or as the isotropic chemical shift (analogous to the chemical shift in the fluid state) *δ*_*i*_, the total anisotropy Δ, and the asymmetry parameter *η*.

The biological specimens generally exhibit spectra with broader linewidths than those of the synthetic compounds, such that the small isotropic chemical shift difference between PO_4_^−3^ and HPO_4_^−2^ prevents these resonances to be separately resolvable. For these spectra, the ssNake program fit the sideband pattern to two CSTs, putatively PO_4_^−3^ and HPO_4_^−2^. The isotropic (liquid-state equivalent) chemical shift *δ*_*i*_ is given by:

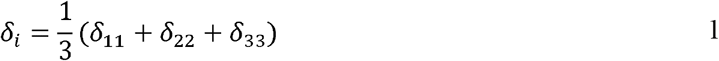

The total anisotropy Δ, a measure of the overall width of the chemical shift powder pattern, can be calculated as:

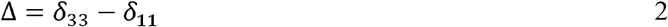

This quantity provides an estimate of the extent to which the point symmetry of the crystal lattice at the nucleus deviates from ideal tetrahedral geometry. When the total anisotropy is zero, the spectral line appears sharp. The asymmetry parameter *η*, which quantifies the deviation of the chemical shift tensor from axial symmetry, is given by:

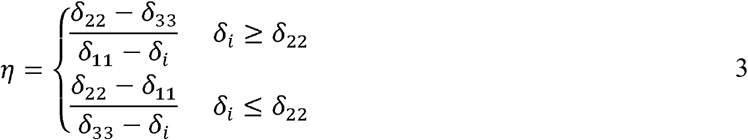

Centerband peak areas were determined by integration to quantify signal intensity *s*(*t*) for extracting CP and rotating frame spin-lattice relaxation (*T*_1*ρ*_) time constants as a function of CP contact time *t*.

The signal was fit to Eq. 4 using a nonlinear least squares approach:

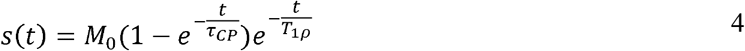

The analysis models the signal as a composite of contributions from both PO_4_^−3^ and HPO_4_^−2^ ions, without attempting to resolve them separately. The rationale behind this model for the time dependence of the ^31^P signal is that the first exponential term describes the buildup of magnetization via rapid ^1^H to ^31^P polarization transfer, while the second exponential term accounts for signal decay due to relaxation in the proton rotating frame. Additional details are provided in the discussion section.

## 3 Results

### 3.1 ^31^P MR spectra

^31^P MAS spectra of the synthetic calcium phosphates and ^1^H-^31^P CPMAS spectra biological specimens are shown in Fig. 1. Because of the high point symmetry of the unprotonated phosphate in hydroxyapatite (Fig. 1(A)), its chemical shift anisotropy is small, and the spinning sidebands are weak. Its centerband (isotropic chemical shift) occurs at 3.1 ppm. There is no indication of other types of phosphate ion in the spectrum. In contrast, monetite (Fig. 1(B)) and brushite (Fig. 1(C)) contain a single acidic phosphate HPO_4_^−2^ and no PO _4_^−3^. The lower symmetry of the HPO_4_^−2^ ion in these compounds results in a large chemical shift anisotropy and a prominent sideband pattern. Their isotropic shifts occur upfield from hydroxyapatite.

The biological specimens exhibit broader spectral linewidths than the synthetic materials. The bone, plaque and calcified valve spectra (Fig. 1(D-F)) resemble that of hydroxyapatite and have isotropic shifts of 3.1 ppm, but with somewhat more prominent sideband patterns similar to monetite and brushite. This suggests that the predominant phosphate ion is PO_4_^−3^, and that it is accompanied by a small amount of HPO_4_^−2^ which is contributing to the sidebands.

The mouse aorta spectrum is distinctly different from those of the other biological specimens in that the sidebands are more prominent, suggesting a higher level of HPO_4_^−2^ content (Fig. 2(G)). The lower spin rate and SNR make visual comparison with the other spectra difficult. However, the analyses below reveal distinct differences from the other biological specimens.

**Figure 2.**
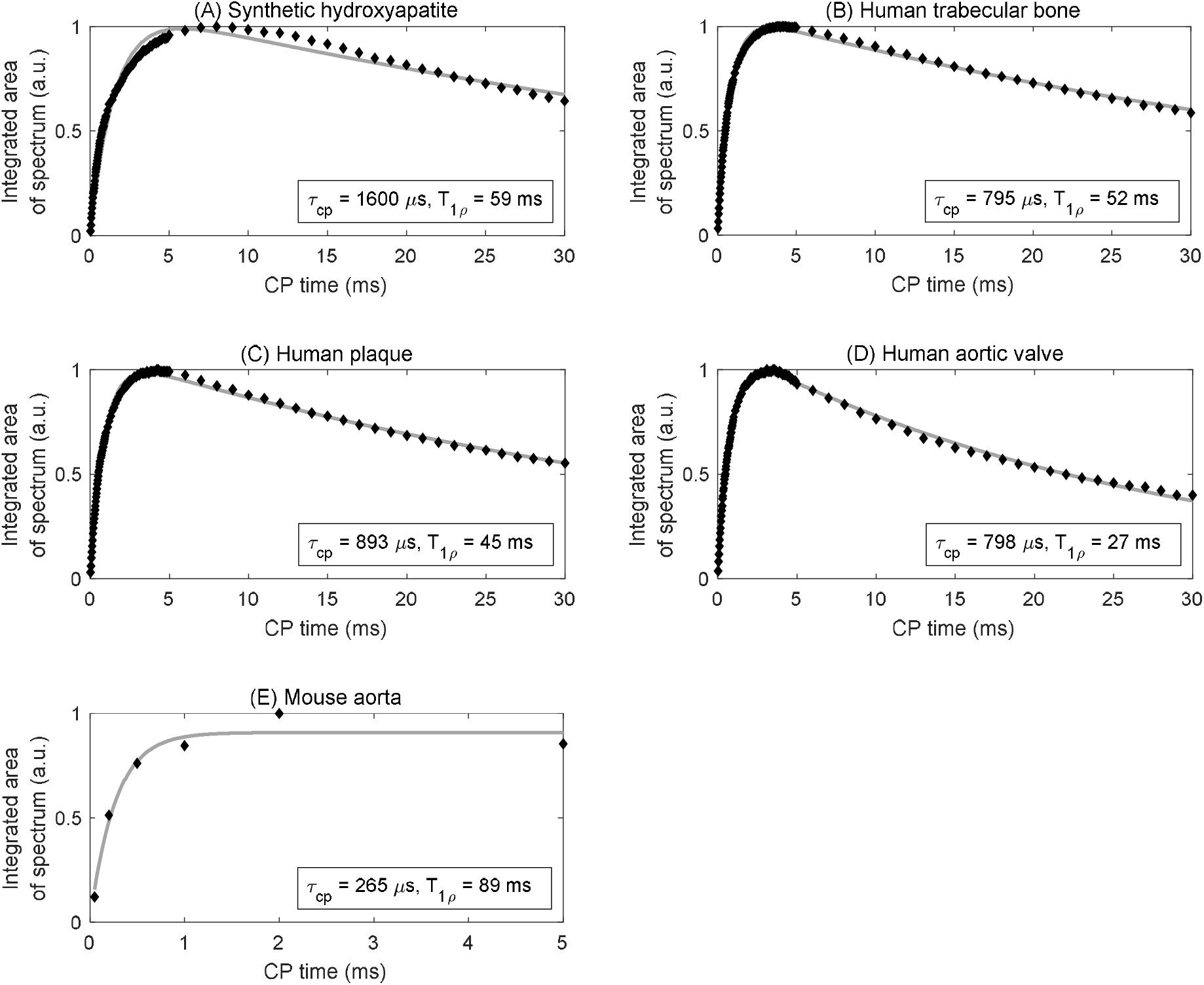
Curve fitting (solid grey line) of integrated area under the centerband (discrete points) provides an estimate of the CP time constant *τ* _*CP*_ and rotating frame spin-lattice relaxation time constant *T*_1*ρ*_ for various samples: (A) synthetic hydroxyapatite, (B) human trabecular bone, (C) human plaque, (D) human aortic valve, and (E) mouse aorta.

### 3.2 Time constants

Signal curves as a function of CP time of selected specimens are shown in Fig. 2, and the corresponding numerical results of the fits are summarized in Table 1. The time axis for the mouse aorta spectrum is limited to 5 ms because of challenges with spinning stability; this *T*_1*ρ*_ value therefore has higher uncertainty. All fits converged successfully.

**Table 1.** Time constants derived from the double-exponential model of CPMAS signal evolution.

| Specimen | $\tau_{CP}(\mu s)^*$ | $T_{1\rho}(ms)^*$ | R-squared |
| --- | --- | --- | --- |
| Hydroxyapatite | $1600 \pm 77$ | $59 \pm 7$ | 0.957 |
| Human trabecular bone | $795 \pm 14$ | $52 \pm 2$ | 0.997 |
| Human plaque | $893 \pm 22$ | $45 \pm 2$ | 0.995 |
| Human aortic valve #1 | $820 \pm 25$ | $56 \pm 4$ | 0.998 |
| Human aortic valve #2 | $733 \pm 21$ | $81 \pm 12$ | 0.998 |
| Human aortic valve #3 | $760 \pm 30$ | $22 \pm 1$ | 0.989 |
| Human aortic valve #4 | $684 \pm 10$ | $36 \pm 1$ | 0.997 |
| Human aortic valve #5 | $798 \pm 10$ | $27 \pm 1$ | 0.997 |
| Mouse aorta** | $265 \pm 133$ | $89 \pm 50$ | 0.974 |

\* Values are mean ± variance. The variance indicates 95% confidence bounds computed using the inverse of Student's t cumulative distribution function and the estimated covariance matrix of the coefficient estimates. \*\*Reduced accuracy due to low SNR and insufficient measurement time for $T_{1\rho}$ quantification.

Hydroxyapatite, containing only unprotonated phosphate, was found to have the longest CP time constant at 1,600 *μ*_*S*_. For comparison, brushite, containing only protonated phosphate, was found in a previous study (43) to have a CP time constant of 140 *μ*_*S*_. The biological specimens in this study fall in between these two extremes. The human specimens tend to be in the range of 700 – 900 *μ*_*S*_, whereas the mouse aorta specimen has a much shorter CP time constant of about 270 *μ*_*S*_, much closer to that of brushite.

### 3.3 Herzfeld-Berger chemical shift tensor

Chemical shift parameters based on Herzfeld-Berger analysis of the spinning sideband patterns are reported in Table 2. The isotropic chemical shifts *δ*_*i*_ are estimated to be accurate to ± 0.2 ppm, and the chemical shift anisotropies Δ are estimated to be accurate to ±2 ppm. The asymmetry parameters *η* are estimated to be accurate to ±0.05. The sideband patterns of the synthetic compounds were fit to a single CST, whereas those of the biological specimens were fit to two CSTs, permitting the separation of unprotonated from protonated phosphates.

The ^31^P chemical shift anisotropy of hydroxyapatite (Δ = 19 ppm) is small as expected because PO _4_^−3^ in hydroxyapatite is located at site of high tetrahedral symmetry. Brushite, which contains a single protonated phosphate ion HPO_4_^−2^, possesses a lower point symmetry, and therefore a larger chemical shift anisotropy (Δ = 125 ppm) than hydroxyapatite. The isotropic shift *δ*_*i*_ of brushite is shifted upfield (less positive) from hydroxyapatite, similar to the upfield shift with decreasing pH (increased protonation) found in aqueous orthophosphates (51, 52). The large value of *η* from brushite indicates a nearly equidistant separation of the principal values of the tensor, and a highly symmetric powder pattern. The *δ*_*i*_ of monetite (CaHPO _4_, the anhydrous form of brushite), lies even further upfield.

Distinct from the “pure” environment of PO_4_^−3^ or HPO_4_^−2^ in the synthetic salts, biological calcium phosphates are mixed crystals containing both types of phosphate ions, as established by indirect wet chemical evidence (53, 54). Biological calcium phosphates also contain miscellaneous crystal lattice defects, vacancies and substitutions. The high surface area of biological calcium phosphate nanocrystals also substantially increases the microscopic heterogeneity of phosphate chemical environments. All of these factors increase the ^31^P spectral linewidths as readily apparent from the spectra in Fig. 1(D-G). Therefore, the analysis of the biological specimens parametrized the spectra as containing contributions from two chemical shift tensors representing PO_4_ ^−3^ and HPO_4_^−2^. Because HPO_4_^−2^ is the minor component, and because the PO_4_^−3^ and HPO_4_^−2^ spectra overlap almost perfectly, a lower accuracy is expected in the HPO_4_^−2^ parameters resulting from the fit. The isotropic chemical shift *δ*_*I*_ calculated from the first tensor of human trabecular bone, plaque and human aortic valves are all at 3.2 ppm (±0.2 ppm), which can be assigned to unprotonated phosphate based on its similarity to the hydroxyapatite isotropic shift. The chemical shift anisotropies Δ calculated for the first tensor of the human specimens are somewhat larger than that of hydroxyapatite. The larger values are likely a reflection of the disordered nature of biological calcium phosphate crystals. The second tensor exhibits isotropic chemical shifts also at 3.2 ppm (±0.2 ppm), except for human aortic valve specimen #2 for which *σ*_*i*_ = 1.4 ppm. The chemical shift anisotropy derived from the second tensor of the human specimens is increased as compared to those of the first tensor, suggestive of some level of “brushite-like” ions in the mineral crystals. The mouse aorta specimen exhibits a spectrum with considerable differences from the other biological specimens. One of the isotropic chemical shifts of mouse aorta lie substantially upfield (*σ*_*i*_ = − 5.2 ppm), implying a higher HPO_4_^−2^ content of the crystals. This was further supported by the increased sideband intensities in Fig. 1(G). Furthermore, an earlier study reported HPO_4_ ^−2^ anisotropy Δ = 116 ppm and *η* = 1 of a few bone specimens from chick embryo, adult chicken, calf embryo, adult calf, rabbit embryo and adult rabbit (43), which are consistently close to values of brushite.

The calculated chemical shift tensors can be used to generate simulated spinning patterns that closely resemble the original spinning patterns, as shown in Fig. 3. For biological specimens characterized by two tensors, the composite pattern is the sum of two simulated patterns. The pattern with a stronger centerband and resonance closer to 3.1 ppm (green traces in Fig. 3(C-F)) is associated with unprotonated phosphates, whereas the pattern with a weaker and broader centerband and resonance shifted upfield (less positive, blue traces in Fig. 3(C-F)) is associated with protonated phosphates. Fitting the mouse spectrum to one tensor gives a reasonable result. The spectrum is too noisy to fit two tensors with reasonable confidence. However, mouse spectrum implies the presence of freshly deposited mineral crystals as opposed to entirely mature hydroxyapatite crystals in the calcification of mouse aorta.

**Figure 3.**
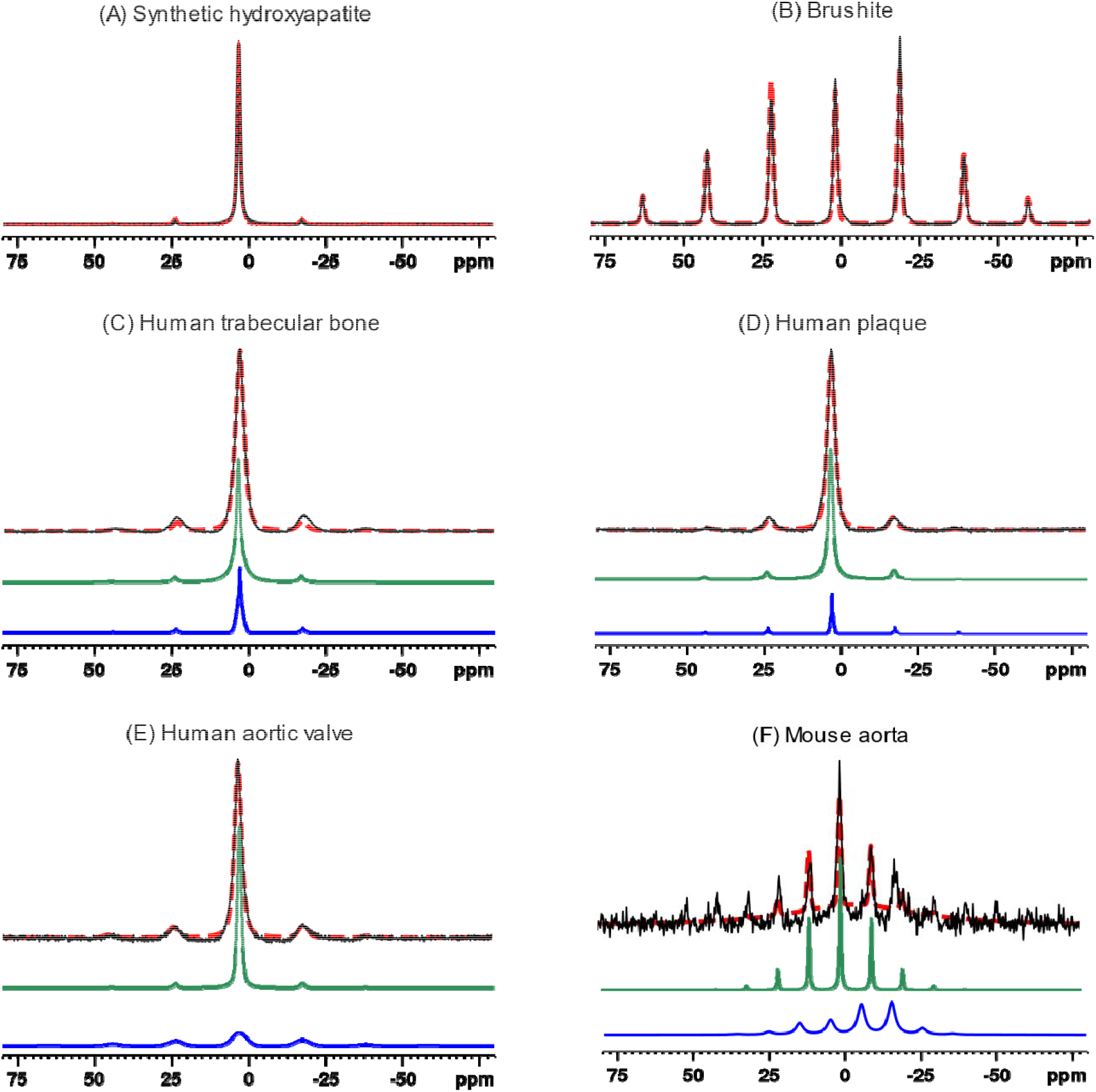
^31^P MAS experimental spectra (solid black traces) of synthetic calcium phosphate compounds and biological specimens, and corresponding simulated spectra (red dashed traces) based on Herfeld-Berger chemical shift tensor analysis. The synthetic compound spectra were fit to a single tensor, whereas biological specimens were fit to two tensors (individual simulated tensor spectra displayed as green and blue traces). The latter composite simulated spectra (red dashed traces) are the sums of the spectra from the two components. (A) Synthetic hydroxyapatite, (B) Brushite, Human trabecular bone, (D) Human plaque, (E) Human aortic valve, (F) Mouse aorta.

## 4 Discussion

The CP rate depends strongly on the direct dipole-dipole coupling between ^1^H and ^31^P nuclei. This rate varies as the inverse sixth power of the internuclear distance and can therefore be used to discriminate between calcium phosphates with close vs. more distant ^31^P–^1^H spacing, as well as deduce general information reflective of the crystal geometry about the phosphate ion. *τ*_*CP*_ also depends on the number of coupled spins. ^31^P in HPO_4_^−2^ transfers magnetization with ^1^H at a rapid rate because the participating nuclei are within the same ion and are in close proximity. At the other end of the scale, PO_4_^−3^ in hydroxyapatite transfers magnetization far more slowly because the closest protons are located in other, more distant, ions (e.g., OH^−^ or HPO_4_^−2^) in the crystal lattice. In this case, ^31^P signals from nuclei separated from protons by more than a few tens of a nm, such as the distance between phosphate ions in calcium phosphate crystals and the protons in surrounding tissue and matrix, are fully suppressed.

T_1ρ_ is the relaxation time constant for energy exchange between nuclear spins and the lattice, measured in the rotating frame of reference. It is analogous to the more familiar spin-lattice relaxation time constant T_1_. T_1_ is sensitive to molecular motions with correlation times comparable to the inverse of the Larmor frequency. For example, for a proton Larmor frequency of 600 MHz (B_0_ ~ 14 T), the proton T_1_ is determined by molecular motion correlation times on the order of ns. A substantial degree of molecular motion (molecular tumbling or diffusion, chemical exchange, or even spin exchange) occurring at this rate will contribute to a shortening in the T_1_ value. Because T_1ρ_ is measured in the rotating reference frame with respect to a spinlock RF field on the order of millitesla (ν_1_ ~ kHz) rather than Tesla (ν_0_ ~ tens to hundreds of MHz), it is sensitive to relatively slow molecular motions (e.g., motions of macromolecules, or slow spin exchange characteristic of substances with dilute, widely spaced, protons).

Spin-lattice relaxation of either type (T_1ρ_ or T_1_) is mediated by the time-dependence (e.g., vibrations and motions of surrounding atoms and molecules) of the spin-spin coupling interaction. Since protons have a much stronger magnetic moment than phosphorus nuclei, they play the dominant role in spin-lattice relaxation. ^31^P nuclei therefore tend to undergo spin-lattice relaxation via spin-spin coupling to the proton “reservoir” (the collective assembly of protons in the crystal lattice, water molecules, bone matrix, etc.). Therefore, variations in T_1*ρ*_ may be indicative of water content, presence of protons in the crystal lattice, and similar aspects of the chemical environment. However, there are no clear trends in the T_1*ρ*_ data for the specimens studied here.

The goal of this study was to provide fundamental insights into the structure of calcified cardiovascular tissues but translational parallels that merit exploration may exist. Cardiovascular MRI in humans is being increasingly explored on 0.5T commercial systems (55, 56) and the relaxation frequency governing T_1_ exchange at this field is not that different from T_1ρ_ at 14T (~36x with the 55 kHz B_1_ field used here). Moreover, portable MRI systems (Hyperfine Inc) operating in the low milli-Tesla range have been FDA-approved for point-of-care neuroimaging and T_1_ of carotid plaque on these systems would involve a relaxation frequency very similar to T_1ρ_ at 14T. These low field systems are based on proton MRI, and the lower polarization of ^31^P would undoubtedly be challenging. However, hyperpolarization of ^31^P in aqueous solution has been described (57) and may be able to mitigate the low SNR of organic phosphates in atherosclerotic plaque. While spatial resolution and SNR are indeed limited in low-field MRI, some valuable insights may be able to be obtained by comparing *in vivo* T_1_ measurements on low field systems with T_1ρ_ measurements *ex vivo* at 14T.

The spectra of all the human specimens resemble that of hydroxyapatite, except the human specimens exhibit more prominent sidebands, as does that of the mouse aorta. As found in earlier studies of animal bone, this is generally the case for mature bone mineral, which is primarily apatitic with a small content of HPO_4_^−2^ that contributes to the sidebands (41, 43). In contrast, calcification in the ApoE-deficient mouse resembled that of immature or embryonic bone. Moreover, calcified mouse aorta exhibits much shorter CP time constants, implying a higher content of HPO_4_^−2^ in the calcium phosphate crystals. Prior studies detected the presence of microcalcifications (1 μm – 30 μm) in the aorta of ApoE-deficient mice fed a high cholesterol diet for as early as 12 weeks starting at 8-10 weeks of age (58, 59) using molecular imaging and microscopy. Despite the age of the ApoE-deficient mouse (60 weeks age) in this study, its calcification did not resemble that observed in vascular calcification in humans. This result could be because calcification in humans may evolve over many years, far longer than the 60 weeks in the mice. Therefore, the use of mouse models for human calcified tissues should be viewed with the caveat that there may be fundamental differences between human and mouse calcification due to the very different time scales for the deposition and remodeling of the calcium phosphate crystals. This may in part explain the discrepancy in the response to statin therapy, which has been effective in ApoE-deficient mice (60–62) but exhibited controversial effects in human patients with atherosclerosis plaques (63–68). Calcification in the aged ApoE-deficient mouse thus likely models early preclinical calcification in human aortic valves (69), but not the mature pattern of calcification seen in patients with established aortic stenosis.

Previous study (43) examining the spectrum of Ca(H_2_PO_4_)_2_ · H_2_ O has ruled out the possibility of the sideband patterns observed in biological calcium phosphates being associated with the protonated phosphate ion H_2_PO_4_^−^. The chemical shift anisotropy of Ca(H_2_PO_4_)_2_ · H_2_ O is much smaller than those of the biological specimens. Additionally, it is unlikely that there is a significant H_2_PO_4_^−^ content in biological calcium phosphates because an unusually acidic cellular environment would be required to maintain a substantial amount of H_2_PO_4_^−^ *in vivo*.

The study is limited by the small number of specimens studied, and the inability to obtain enough calcified mouse aorta to acquire high SNR spectra in a reasonable amount of time. It was assumed that only two phosphate ions (PO _4_^−3^ and HPO_4_^−2^) contribute to the spectra. In principle, each sideband pattern is affected by not only the ^31^P chemical shift anisotropy, but also ^1^H-^31^P and ^31^P-^31^P dipolar interactions. However, these secondary effects are expected to be relatively minor under conditions of high magnetic field strength and 5 kHz magic angle spinning.

## 5 Conclusions

Solid state MR spectroscopy adds another dimension to the study of pathologic calcification. It provides a means to identify the presence of various phosphate ions in the calcium phosphate mineral. Human trabecular bone, calcified atherosclerotic plaque, and calcified aortic valve specimens were found to have similar CPMAS spectra, whereas the spectra of experimentally induced calcified mouse aortic calcification are somewhat different and suggestive of immature, recently deposited bone. Although solid state MR spectroscopy has been used to study bone in several contexts, it has rarely been applied to pathological ectopic calcification. While this methodology is not directly applicable to clinical use, it could be an important complement to basic studies of pathological calcification in atherosclerosis and related calcific disorders.

## Data Availability

All data produced in the present study are available upon reasonable request to the authors.

## Data availability statement

Data supporting this study are available from the corresponding author upon request.

## Author contributions

JLA, DES, and EA conceived the study. EA and BFM provided and prepared the mouse specimens, JLA provided the human bone and plaque specimens, and DES provided the human aortic valve specimens. CTF acquired magnetic resonance spectroscopy data. JY analyzed the data and drafted the manuscript, with AK assisting in the Herzfeld–Berger analysis. All authors reviewed and approved the final version of the manuscript.

